# A CLOSER LOOK AT INDIRECT CAUSES OF DEATH AFTER HURRICANE MARIA USING A SEMIPARAMETRIC MODEL

**DOI:** 10.1101/2023.01.03.23284158

**Authors:** Oscar Lugo, Roberto Rivera

## Abstract

The Covid-19 pandemic as well as other recent natural emergencies have put the spotlight on emergency planning. One important aspect is that natural disasters or emergencies often lead to indirect deaths and studying the behavior of indirect deaths during emergencies can guide emergency planning. While many studies have implied a large number of indirect deaths in Puerto Rico due to Hurricane Maria; the specific causes of these deaths have not being carefully studied. In this paper, we use a semiparametric model and mortality data to evaluate cause of death trends. Our model adjusts for cause of death effect potentially varying over time while also inferring on how long excess deaths occurred. From September 2017 to March 2018, after adjusting for intra-annual variability and population displacement, we find evidence of significant excess deaths due to Alzheimer’s/Parkinson, heart disease, sepsis, diabetes, renal failure, and pneumonia & influenza. In contrast, for the same time period we find no evidence of significant excess deaths due to cancer, hypertension, respiratory diseases, cerebrovascular disease, suicide, homicide, falling accidents and traffic accidents.

## 1 INTRODUCTION

On September 20 2017, Hurricane Maria struck Puerto Rico with 155 miles per hour sustained winds and wind gusts of up to 190 mph (Pasch et al., 2018). Torrential rains of up to 40 inches of water accompanied the winds (Hu and Smith, 2018), which resulted in large floods on the entire island. The hurricane destroyed the island’s electricity grid, leaving 3.4 million inhabitants without electricity (Lu and Alcantara, 2020). In addition, infrastructure was affected: aqueducts and sewers, roads, bridges, ports, airports, cell phone towers, and gas stations were left without fuel for vehicles and generators (Ficek, 2018). It is estimated that the effects of the hurricane caused monetary damages in Puerto Rico for more than the US $82 billion (Álvarez Febles and Félix, 2020). Also, Maria destroyed around 80% of the value of the island’s crops, representing a loss of $780 million in agricultural yield (Rodríguez Cruz and Niles, 2018).

Indirect causes of death have been recognized as an important consequence of national emergencies. For example, damage to the transportation network during an earthquake can affect access to medicine and health services generating serious or even fatal repercussions for affected people (Geronimus et al., 2006; Haentjens et al., 2010; WHO, 2018a). However, the different interpretations of guidelines and varied training of personnel determining the cause of death hinder mortality counts (German et al., 2001). Several studies have looked into excess deaths due to Hurricane Maria (Santos-Lozada and Howard, 2017; Rivera and Rolke, 2018; Kishore et al., 2018; Santos-Burgoa et al., 2018; Rivera and Rolke, 2019; Cruz-Cano and Mead, 2019). While Cruz-Cano and Mead (2019) assessed specific indirect causes of death after Hurricane Maria, it only did so until the end of October 2017. Moreover, the authors limited their analysis to a few causes (heart disease, other causes, diabetes, Alzheimer’s disease, and septicemia), and did not account for population displacement, possibly leading to an underestimation of excess deaths. Importantly, the duration of Hurricane Maria’s excess mortality effect may have differed by cause of death, and modeling this can guide the development of emergency management planning.

For future emergency planning and preparedness purposes, it is important to predict fatalities related to the state of emergency, whether the deaths are directly or indirectly related to the event (Brunkard et al., 2008; Ragan et al., 2008; Rivera et al., 2020; Rosenbaum et al., 2021) and excess mortality is a useful method of assessing deaths due to an emergency (Hiam et al., 2017). This research focuses on building a semi-parametric Poisson model that allows estimating the deaths caused by a state of emergency incorporating in the model the intra-annual variability, the population displacement, and causes of death due to non-communicable diseases, and other causes indirect. At the same time, the model captures the duration of the effect on mortality of an emergency according to each death cause.

## 2 A Semiparametric Model

We propose a semiparametric Poisson regression model Wood (2017) to estimate whether some deaths due to some causes increased after the passage of Hurricane Maria in Puerto Rico while controlling for the temporal variation in the mortality rate of the causes of death and the population size. Population displacement plays an important role, which can be directly influenced by economic, social, labor, academic, climatic variations, and other factors (Neyra, 2017; Acosta et al., 2020).

Additionally, index functions will be deployed in the model to evaluate if there are effects of changes in the behavior of certain leading causes of death including some Non-Communicable Diseases (NCDs) in different post-emergency periods.

Let *D*_*t*_ = number of deaths at time index *t, N*_*t*_ = population size at time *t*. We assume that *D*_*t*_ follows a Poisson distribution. For *i* = 0, …, I, we assign p_*i,t*_ as an indicator of the period *i* for time *t*. Specifically, *i* = 0 represents the pre-emergency period, *i* = 1 is a given period after the emergency, and so on. For *j* = 1, … J, we consider c_*j,t*_ as an indicator of cause of death *j* at time *t*. On the other hand, we define day_*t*_ = day of the year, and year_*t*_ = a categorical variable that captures the mortality trend through the years. We define the following generalized additive semiparametric Poisson regression model with interaction (Ruppert et al., 2003), 

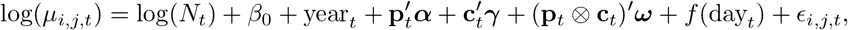

where *μ*_*i,j,t*_ = *E*(*D*_*t*_ *t*, 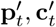, *N*_*t*_, day_*t*_, year_*t*_). We define 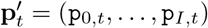 and ***α*** = (*α*_0_, …, *α*_*I*_)^*′*^ denotes the coefficients of the model for the *I* pre and post-emergency periods. Similarly, 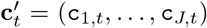 and ***γ*** = (*γ*_1_, *γ*_2_, …, *γ*_*J*_)^*′*^ denotes the coefficients of the model for the *J* causes of death. While (**p**_*t*_⊗ **c**_*t*_)^*′*^ = (p_0,*t*_c_1,*t*_, …, p_*I,t*_c_*J,t*_) is the Kronecker product between vectors 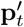 and 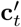 represents the interaction between the *I* pre or post-emergency periods and the *J* causes of death at time *t* and ***ω*** = (*ω*_11_, …, *ω*_*MQ*_)^*′*^ represents the coefficients of the interaction. On the other hand, the smooth function *f* (day_*t*_) captures within-year variation in deaths while *β*_0_ is the contrast of the smooth function and the explanatory variables, and *ϵ*_*i,j,t*_ contains the variability not explained by the other terms. For post-emergency periods, p_*i,t*_ = 1 and the interaction coefficient make it possible to adapt the effect of the post-emergency period for cause *j*. The coefficients of the model (1) are estimated by penalized likelihood maximization, taking into account the population displacement caused by a state of emergency while the smoothing penalty parameters are determined by the restricted maximum likelihood (REML). Model (1) can be used to estimate excess death of cause of death *j* during period *i* post-emergency of index time *t* through the difference between the estimation of the model with p_*i,t*_ = 1, c_*j,t*_ = 1 and p_*i,t*_c_*j,t*_ = 1 versus the estimated model with p_*i,t*_ = 0, c_*j,t*_ = 1 and p_*i,t*_c_*j,t*_ = 0. For *i* ≥ 1 and *j* ≥ 1 we obtain:

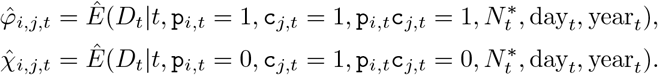

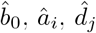 and *ê*_*i,j*_ estimate *β*_0_, *α*_*i*_, *γ*_*j*_ and *ω*_*ij*_ respectively. Also, notice that 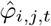 uses 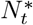, a population size unaltered by post-emergency displacement because *â*_*i*_ and *ê*_*ij*_ are already considering the impact of population displacement. Using it again for 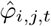 would imply accounting for the effect twice. This way,

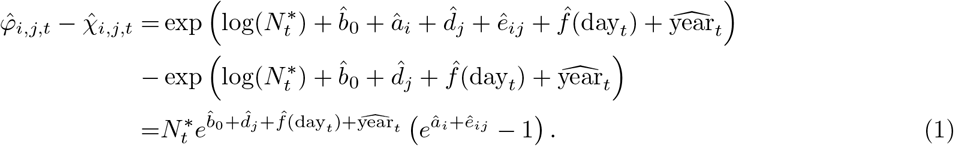

The expression 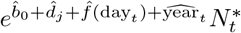 in (1) represents the number of deaths typically seen per and 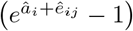 represents percentage changes during the post-emergency periods adjusting for the interaction between the *J* causes of death and the post-emergency periods. Equation (1) is the maximum likelihood estimator for the excess deaths expected from the *J* causes of death in post-emergency period *i*. Using equation (1) the accumulated excess of the *J* causes of death is,

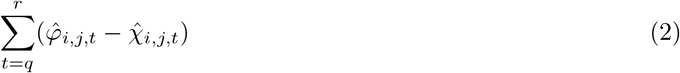

for any period that begins at the index *q* and ends at *r*.

In general terms, model (1) allows statistical inference of the duration of the effect in the *I* post-emergency periods, the effect on the *J* causes of death during a state of emergency, and the effect caused by the interaction between the *I* post-emergency periods and the *J* causes of death in the population displacement.

## 3 Results

### 3.1 Data

The data used during this study correspond to the deaths reported daily by the Puerto Rico Demographic and Vital Statistics Registry between January 1, 2014 to December 31, 2018 and includes a total of 148,490 records. This work is exempt from requiring human subjects board review. Death records without autopsy are processed in the software “Automatic Classification of Medical Entry”(ACME), this program selects the underlying cause of death (Variables Coding and ACME Description) using the information based on the codes of the International Classification of Diseases (ICD-10 (2016)) on all medical conditions reported for death along with information on the reported relationship between the various conditions (Lu, 2003).

### 3.2 Population displacement

Demographic data indicates that Puerto Rico registers a high migratory movement, due to the economic crisis it has faced since 2006 (DeWaard et al., 2020; Castro-Prieto et al., 2017; Abel and Deitz, 2014; Rivera, 2016). After the passage of Hurricane Maria in Puerto Rico, lack of water or water service (DeWaard et al., 2020), and a health service crisis (Panditharatne, 2018; Watkins et al., 2020; Glassman, 2019) lead to an increase in population displacement, which can permanently alter the demographic composition of the affected regions (Acosta et al., 2020).

Like Rivera and Rolke (2019) we estimate population displacement through annual estimates of the resident population of Puerto Rico according to the data US Census Bureau adjusted for passenger movement. Usually, the movement passenger is a biased indirect variable of resident migration since it includes the visitor seasonal movements (Velazquez-Estrada et al., 2018), and tourism has been one of the few growing sectors in the Puerto Rican economy (Rivera, 2016). However, a dramatic drop in visitors (Rivera and Rolke, 2019) occurred the months after the arrival of Hurricane Maria. Other methods like the use of disaster maps (Echenique and Melgar, 2018) did not significantly affect our results.

Data from daily death certificates (*D*_*t*_) were combined with population estimates (*N*_*t*_) to produce daily mortality rates (*R*_*t*_);

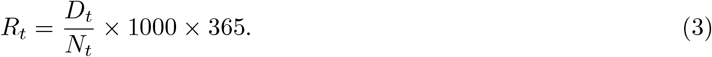

Figure 1 illustrates the daily mortality rate during the years 2014-2018 in Puerto Rico. This graph indicates that the first and last months of each year generally have the highest death rates a pattern commonly seen in other locations (Phillips et al., 2010). On the other hand, Figure 1 displays a drastic spike in mortality rate in the last days of September 2017 and remained high until the beginning of 2018. It is not clear from the figure whether mortality rates increased over the years, yet our model of excess deaths takes the variable `year`_*t*_ can explain this type of temporal fluctuation.

**Figure 1:**
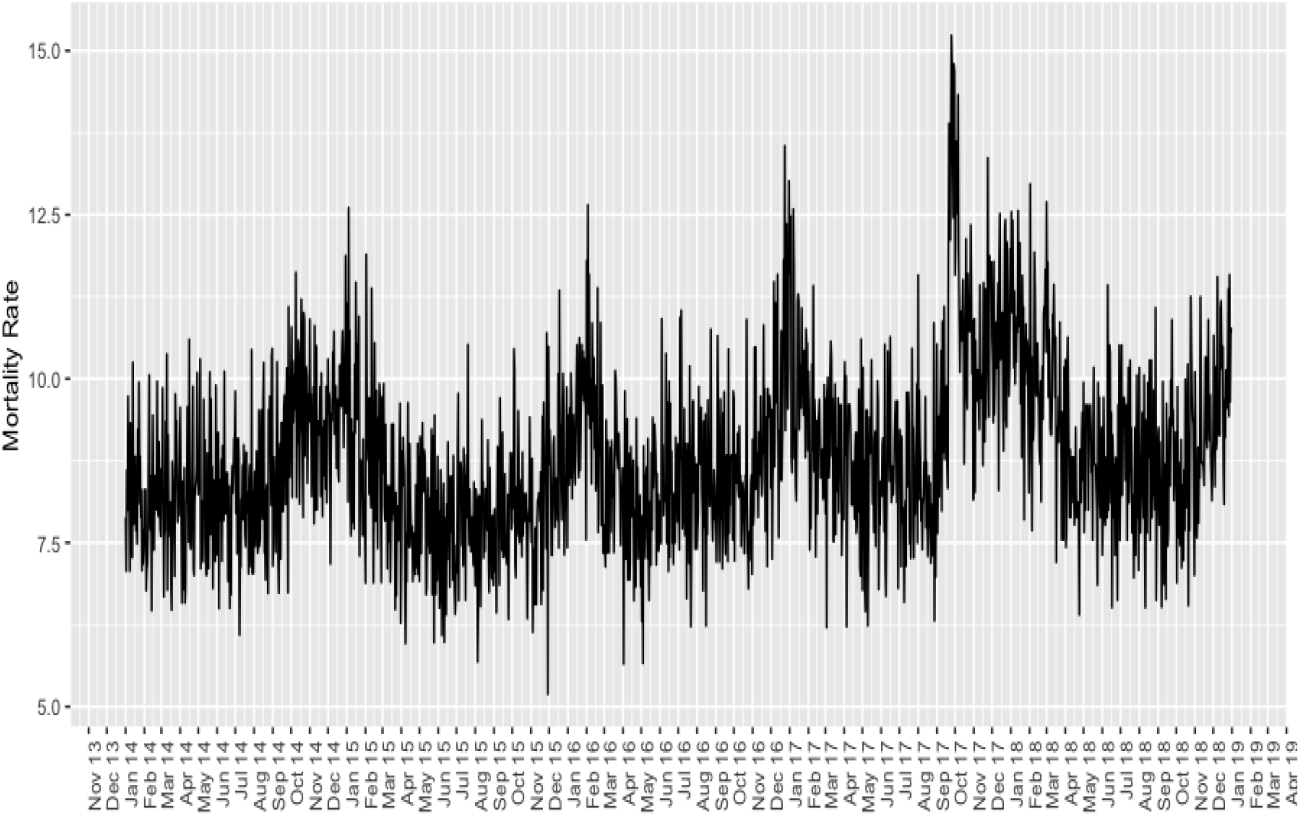
Average daily mortality rate from 2014-2018.

In the preliminary analysis, the models did not indicate over-dispersion, so a Poisson model was used.

### 3.3 Cumulative Excess Deaths by Cause

According to the World Health Organization (WHO, 2018b), the Institute of Statistics of Puerto Rico (Health Department, 2013), and Cruz-Cano and Mead (2019), the main causes of death in Puerto Rico are cancer, hypertension, cardiovascular, kidney, respiratory diseases, and conditions that affect the nervous and endocrine systems. Other causes of death of interest are traffic accidents, homicides, and suicides (Petrosky et al., 2020). In this study, we consider these causes of death along with sepsis, accidents due to any type of fall, and ‘undefined deaths’ to determine if said causes presented an excess of mortality after the natural disaster.

No interannual pattern is observed in the selected causes of death (Figure 2). On the other hand, deaths due to “Other Causes of Death”, “Heart”, “Cancer” and “Diabetes” had the highest average monthly mortality rate, followed by deaths due to “Alzheimer’s or Parkinson’s”, “Respiratory Diseases”, “Hypertension”, “Cerebrovascular”, “Sepsis”, “Pneumonia and Influenza”, “Renal Insufficiency”, “Homicide”, and “Undefined Causes”. Lastly, “Suicide”, “Fall Accidents” and “Traffic” display low mortality rates. We use the class “Other Causes of Death” and the year “2014” as the reference categories for these variables.

**Figure 2:**
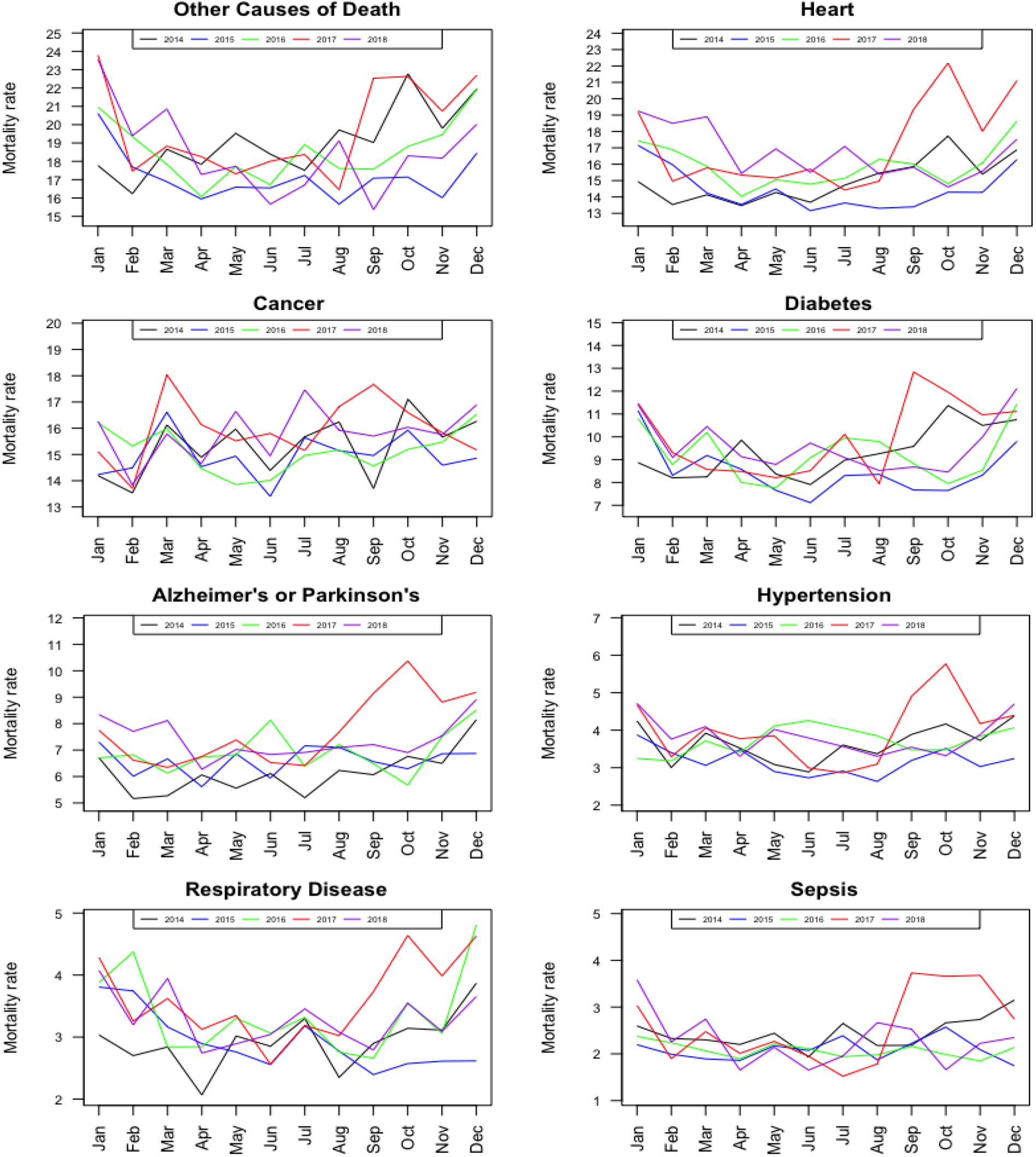
Monthly mortality rates for several causes of death during the years 2014-2018 in Puerto Rico.

Table 1 presents the estimates of excess and cumulative excess deaths for the 14 causes of death according to 7 post-Maria periods and their respective 95% confidence intervals based on model (1).

**Table 1:** Estimates of excess death and cumulative excess death for the different causes of death according to model 1 and their respective 95% confidence intervals.

|  | Other |  | Cancer |  | Heart |  | Diabetes |  | Alzheimer's & Parkinson |  | Hypertension |  |
| --- | --- | --- | --- | --- | --- | --- | --- | --- | --- | --- | --- | --- |
| Period | Est. | 95% CI | Est. | 95% CI | Est. | 95% CI | Est. | 95% CI | Est. | 95% CI | Est. | 95% CI |
| Sep | 27 | (13 , 45) | 21 | (-3 , 51) | 99 | (68 , 132) | 93 | (67 , 122) | 47 | (27 , 71) | 16 | (5 , 32) |
| Oct | 51 | (27 , 78) | 0 | (-41 , 45) | 148 | (99 , 200) | 56 | (21 , 95) | 82 | (50 , 118) | 21 | (2 , 42) |
| Sep - Oct | 78 | (41 , 124) | 21 | (-45 , 98) | 247 | (168 , 332) | 150 | (89 , 217) | 130 | (78 , 189) | 37 | (7 , 74) |
| Nov | 10 | (-9 , 33) | -18 | (-58 , 26) | 40 | (-3 , 88) | 31 | (-1 , 68) | 42 | (13 , 75) | -2 | (-16 , 17) |
| Sep - Nov | 88 | (31 , 157) | 3 | (-103 , 124) | 288 | (164 , 420) | 181 | (88 , 285) | 171 | (90 , 264) | 35 | (-9 , 91) |
| Dic | 6 | (-13 , 30) | -71 | (-110 , -29) | 85 | (38 , 135) | 14 | (-19 , 51) | 35 | (6 , 70) | 6 | (-10 , 27) |
| Sep - Dec | 94 | (17 , 188) | -68 | (-214 , 95) | 372 | (203 , 555) | 195 | (68 , 336) | 207 | (96 , 334) | 41 | (-19 , 118) |
| Jan | 17 | (-3 , 41) | -35 | (-76 , -10) | 45 | (0 , 94) | 26 | (-8 , 63) | 17 | (-11 , 50) | 8 | (-9 , 28) |
| Sep - Jan | 111 | (14 , 229) | -103 | (-290 , -106) | 417 | (203 , 649) | 221 | (60 , 399) | 224 | (86 , 385) | 49 | (-29 , 147) |
| Feb | 7 | (-11 , 29) | -35 | (-73 , -7) | 90 | (46 , 138) | 4 | (-27 , 38) | 29 | (2 , 60) | 18 | (0 , 38) |
| Sep - Feb | 118 | (2 , 259) | -138 | (-363 , 114) | 507 | (249 , 787) | 225 | (33 , 438) | 253 | (88 , 445) | 67 | (-28 , 186) |
| Mar | 11 | (-8 , 34) | -4 | (-45 , 40) | 80 | (36 , 128) | 27 | (-5 , 63) | 31 | (3 , 63) | 9 | (-6 , 29) |
| Sep - Mar | 129 | (-6 , 293) | -142 | (-408 , 154) | 587 | (284 , 916) | 252 | (28 , 501) | 248 | (90 , 508) | 76 | (-35 , 215) |
|  | Respiratory Diseases |  | Cerebrovascular |  | Sepsis |  | Pneumonia & Influenza |  | Renal Insufficiency |  |  |  |
| Period | Est. | 95% CI | Est. | 95% CI | Est. | 95% CI | Est. | 95% CI | Est. | 95% CI |  |  |
| Sep | 13 | (1 , 29) | 1 | (-10 , 15) | 33 | (20 , 51) | 20 | (9 , 36) | 11 | (2 , 24) |  |  |
| Oct | 34 | (13 , 58) | 20 | (-1 , 44) | 35 | (16 , 56) | 23 | (6 , 44) | 42 | (25 , 64) |  |  |
| Sep - Oct | 47 | (14 , 87) | 20 | (-11 , 59) | 68 | (37 , 108) | 44 | (15 , 80) | 53 | (27 , 89) |  |  |
| Nov | 17 | (-2 , 40) | 18 | (-2 , 43) | 35 | (17 , 56) | 24 | (7 , 44) | 22 | (6 , 40) |  |  |
| Sep - Nov | 63 | (12 , 127) | 39 | (-14 , 103) | 103 | (54 , 164) | 67 | (22 , 124) | 75 | (34 , 130) |  |  |
| Dec | 26 | (5 , 51) | -11 | (-29 , 12) | 5 | (-10 , 24) | 35 | (16 , 57) | 28 | (11 , 47) |  |  |
| Sep - Dec | 89 | (17 , 177) | 28 | (-43 , 115) | 108 | (43 , 189) | 102 | (39 , 181) | 103 | (45 , 178) |  |  |
| Jan | 13 | (-5 , 37) | -11 | (-29 , 11) | 29 | (10 , 50) | 49 | (30 , 73) | -8 | (-20 , 7) |  |  |
| Sep - Jan | 103 | (12 , 215) | 17 | (-73 , 126) | 137 | (54 , 239) | 152 | (68 , 254) | 95 | (25 , 185) |  |  |
| Feb | 3 | (-14 , 24) | 10 | (-9 , 32) | 3 | (-10 , 20) | 20 | (4 , 41) | 16 | (2 , 35) |  |  |
| Sep - Feb | 106 | (-2 , 239) | 26 | (-82 , 158) | 140 | (43 , 260) | 172 | (73 , 295) | 111 | (27 , 220) |  |  |
| Mar | 19 | (0 , 42) | 8 | (-11 , 31) | 13 | (-2 , 32) | 22 | (6 , 43) | 18 | (4 , 36) |  |  |
| Sep - Mar | 125 | (-2 , 281) | 34 | (-93 , 189) | 153 | (41 , 292) | 195 | (79 , 338) | 129 | (31 , 257) |  |  |
|  | Homicide |  | Undefined |  | Suicide |  | Falling Accident |  | Traffic Acc |  |  |  |
| Period | Est. | 95% CI | Est. | 95% CI | Est. | 95% CI | Est. | 95% CI | Est. | 95% CI |  |  |
| Sep | -1 | (-8 , 9) | 91 | (60 , 127) | 4 | (-1 , 14) | 1 | (-3 , 9) | -7 | (-10 , -1) |  |  |
| Oct | 11 | (-3 , 29) | 78 | (28 , 131) | 4 | (-4 , 16) | 8 | (-1 , 21) | -6 | (-15 , 6) |  |  |
| Sep - Oct | 9 | (-12 , 39) | 169 | (88 , 259) | 8 | (-4 , 30) | 8 | (-4 , 30) | -14 | (-24 , 5) |  |  |
| Nov | -3 | (-16 , 13) | 31 | (-16 , 80) | 8 | (-1 , 20) | 8 | (-1 , 21) | 12 | (1 , 28) |  |  |
| Sep - Nov | 6 | (-28 , 52) | 200 | (71 , 340) | 16 | (-6 , 50) | 17 | (-5 , 51) | -2 | (-23 , 33) |  |  |
| Dec | 7 | (-7 , 26) | 37 | (-11 , 90) | 1 | (-6 , 13) | 3 | (-5 , 15) | 20 | (7 , 36) |  |  |
| Sep - Dec | 14 | (-35 , 78) | 238 | (59 , 430) | 17 | (-12 , 63) | 19 | (-10 , 66) | 18 | (-16 , 69) |  |  |
| Jan | 22 | (6 , 42) | 70 | (19 , 124) | -7 | (-13 , 2) | 11 | (1 , 24) | 3 | (-7 , 17) |  |  |
| Sep - Jan | 36 | (-29 , 120) | 309 | (78 , 555) | 10 | (-25 , 66) | 30 | (-9 , 90) | 21 | (-23 , 86) |  |  |
| Feb | 7 | (-6 , 25) | 39 | (-5 , 89) | 2 | (-5 , 13) | 0 | (-7 , 12) | 1 | (-8 , 14) |  |  |
| Sep - Feb | 43 | (-35 , 145) | 348 | (73 , 644) | 12 | (-30 , 79) | 31 | (-16 , 102) | 22 | (-31 , 100) |  |  |
| Mar | 3 | (-10 , 20) | 53 | (7 , 105) | 6 | (-2 , 18) | 6 | (-3 , 18) | -4 | (-13 , 8) |  |  |
| Sep - Mar | 46 | (-45 , 166) | 402 | (80 , 750) | 18 | (-32 , 97) | 36 | (-19 , 120) | 18 | (-44 , 107) |  |  |

21 excess deaths during September 20th to 30th, 2017 were estimated for “Cancer”, this period being the only one that presented excess deaths. Consequently, the cumulative total excess death intervals with 95% confidence from October 2017 to March 2018 included zero. For October 2017, 148 excess deaths are estimated for “Heart Diseases”. For the periods from September 20 to 30, December, February, and March, the excess ranged from 80 to 99 deaths. While, from September to March we are 95% confident that (284, 916) excess heart disease deaths occurred. For “Diabetes”, the first two post-Maria periods showed an excess of 93 and 56 deaths, respectively. For the entire post-Maria period, considerable uncertainty is found on cumulative excess deaths: (28, 501). It was found that for “Alzheimer’s or Parkinson’s”, “Pneumonia & Influenza”, “Sepsis” “Renal Insufficiency” and “Undefined” excess deaths during the entire post-Maria period were significant. According to the 95% confidence intervals for causes “Suicide”, “Falling Accidents” and “Traffic”, there is no statistically significant evidence of excess deaths from September 20-30, 2017 to March 2018.

Overall, the causes of death with the largest excess deaths during the seven post-emergency periods were: “Heart” and “Undefined”. It should be emphasized that the excess from November 2017 to January 2018 for “Pneumonia and Influenza” were not necessarily due to Maria as seasonal cases are expected (Paz-Bailey et al., 2020).

Figures 3 and 4 display cumulative excess deaths for all causes; in red is the estimated value while black functions are upper and lower 95%. confidence bounds.

**Figure 3:**
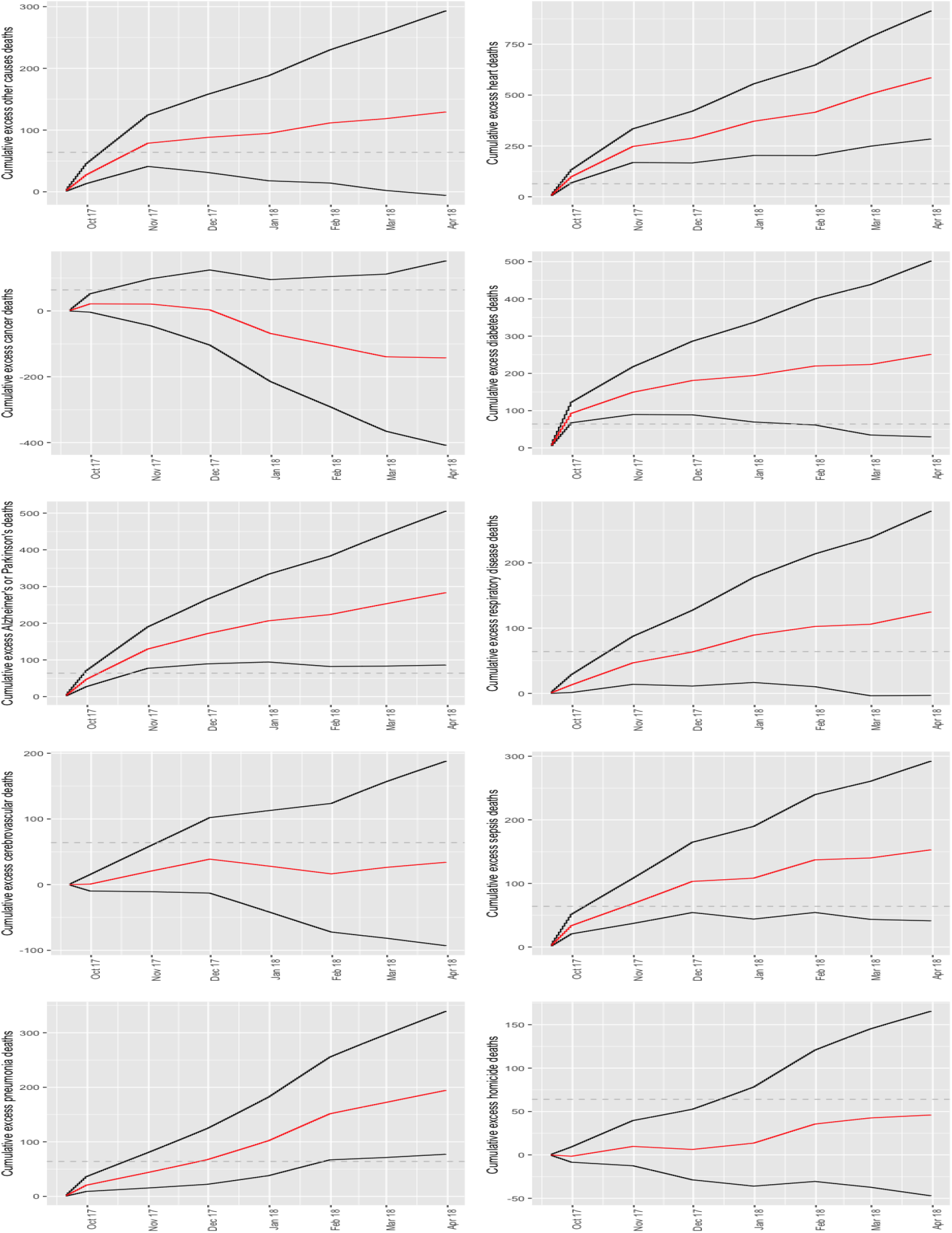
Cumulative excess death estimates from September 20 to March 31, 2018 according to: other causes, heart disease, cancer, diabetes, Alzheimer or Parkinson’s, respiratory disease, cardiovascular disease, sepsis, pneumonia, and homicide. Dashed line represents the 64 deaths authorities initially attributed to Hurricane Maria.

**Figure 4:**
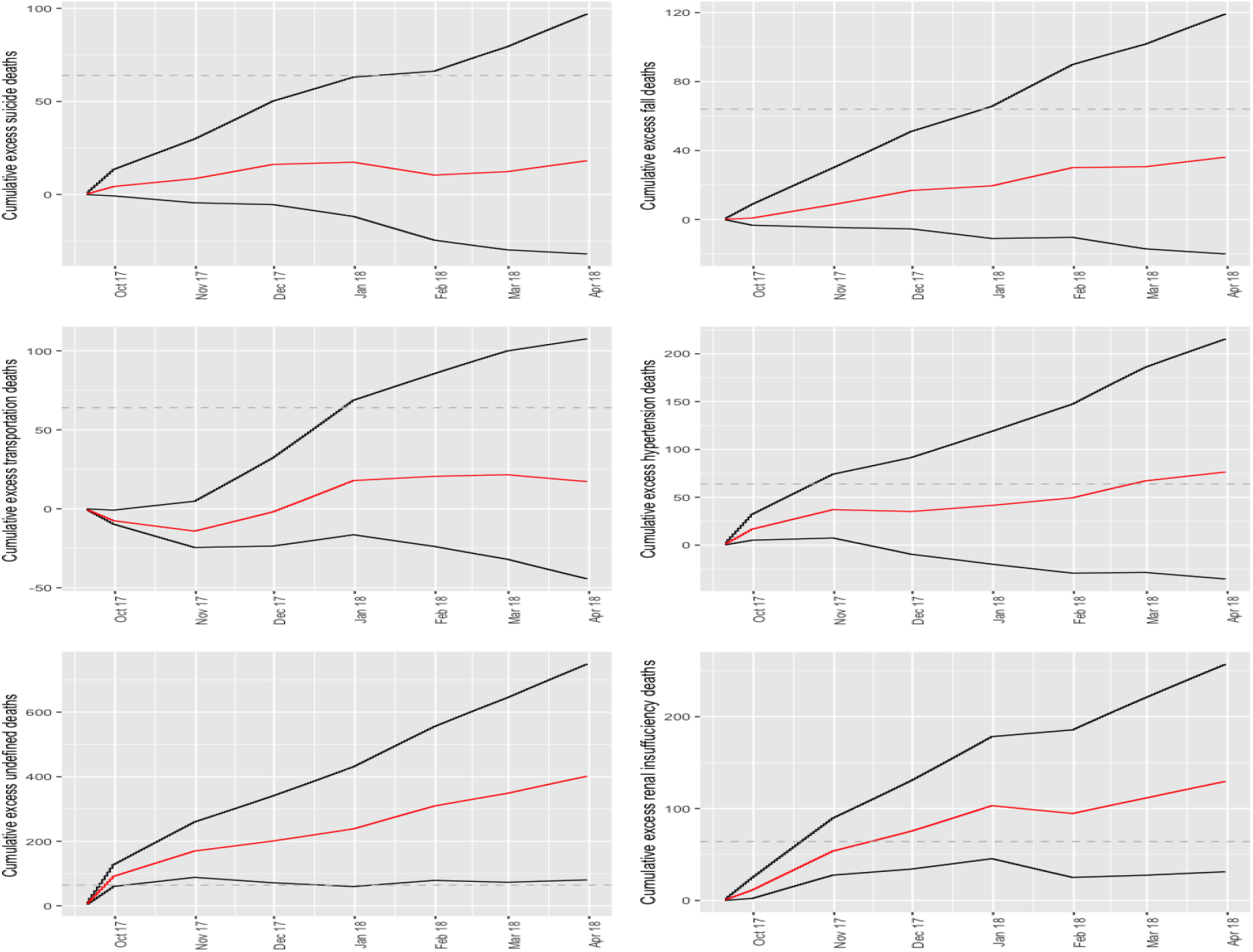
Cumulative excess death estimates from September 20 to March 31, 2018 according to: suicide, fall, traffic accident, hypertension, undefined, and renal insufficiency. Dashed line represents the 64 deaths authorities initially attributed to Hurricane Maria.

## 4 Discussion

This study explores the excess mortality due to non-communicable diseases and other indirect causes during the state of emergency generated after Hurricane Maria’s passage over Puerto Rico. The model makes adjustments for intra-annual variability and population displacement. This allows greater precision when estimating excess mortality. Moreover, our method can infer on how long an emergency leads to an increase in deaths. To achieve our objective, the statistical model from Rivera and Rolke (2019) was extended to account for different causes of deaths and included cause of death and post-emergency interaction terms so that the model can adjust the effect during the post-emergency period *i* for each cause of death.

It is determined that the causes of deaths due to sepsis, and diabetes, presented the highest excess deaths for residents of Puerto Rico during September and October, 2017 compared to the pre-Maria period and relative to “Other Causes of death”. Excess death due to heart disease, Alzheimer’s/Parkinson, pneumonia & influenza and renal failure persisted for months after landfall.

The excess deaths observed from renal insufficiency may be attributed to the dialysis centers being left without electric power and electric generators not being designed to be used for long periods (Michaud and Kates, 2017). On the other hand, an increase in sepsis deaths may be due to health problems caused by a lack of supply of potable water since 20 out of 51 wastewater treatment plants were out of service one month after the arrival of Hurricane Maria (Michaud and Kates, 2017). For its part, the excess mortality from this disease was caused by multiorgan dysfunction syndrome, due to poor access to care in intensive care units and the shortage of drug supply (Driessen et al., 2021). Furthermore, Alzheimer’s excess deaths could be due to people who suffer from this disease tending to live in care centers, and the state of emergency led to a reduction in their medical care, routine treatments (Ruhm, 2021). Difficulty in continuing with adequate nutrition adequate medical care, lack and loss of medicines, and difficulties in monitoring their levels of glucose resulted in an increase in deaths due to diabetes (Quast et al., 2019).

Cruz-Cano and Mead (2019) evaluated some causes of death that were traced and reported by the Department of Health of Puerto Rico but only until October 2017. Our estimates are comparable to theirs, though we study more causes of deaths, find evidence Hurricane Maria lead to excess deaths until March 2018, we find significant renal failure excess deaths and infer there was an increase in deaths where the cause was left undefined.

Estimating excess deaths in Puerto Rico by cause after Hurricane Maria helps us understand the effect of the natural disaster and allows us to assess the weaknesses in the health system to better face these types of events in the future (Perreira et al., 2016). Authorities need tools to reduce the number of deaths due to state emergencies. Some strategies include the use of technology in telemedicine, applications for mobile devices, virtual programs (Hacker et al., 2021) and remote monitoring Bowman et al. (2021); LRM (2020). Taking into account the above, it is expected that these measures will reduce the risk of death in the population and better prepare health professionals for future states of emergency generated by different disasters (e.g. hurricanes, tsunamis, and pandemic).

Competing interests: The author(s) declare none

## Data Availability

Available upon request

